# Neuropsychiatric disorders as risk factors and consequences of COVID-19: A Mendelian randomization study

**DOI:** 10.1101/2021.06.29.21259609

**Authors:** Yong Xiang, Jinghong Qiu, Ruoyu Zhang, Carlos Kwan-Long Chau, Shitao Rao, Hon-Cheong So

**Author notes:** **Correspondence to: Hon-Cheong So**, Lo Kwee-Seong Integrated Biomedical Sciences Building, The Chinese University of Hong Kong, Shatin, Hong Kong.

## Abstract

**Background:** More than 180 million cases of COVID-19 have been reported worldwide. It has been proposed that neuropsychiatric disorders may be risk factors and/or consequences of COVID-19 infection. However, observational studies could be affected by confounding bias.

**Methods:** We performed bi-directional two-sample Mendelian randomization (MR) analysis to evaluate causal relationships between liability to COVID-19 (and severe/critical infection) and a wide range of neuropsychiatric disorders or traits. We employed GWAS summary statistics from the COVID-19 Host Genetics Initiative. A variety of MR methods including those accounting for horizontal pleiotropy were employed.

**Results:** Overall, we observed evidence that liability to COVID-19 or severe infection may be causally associated with higher risks of post-traumatic stress disorder (PTSD), bipolar disorder (BD) (especially BD II), schizophrenia (SCZ), attention deficit hyperactivity disorder (ADHD) and suicidal thought (ST) when compared to the general population. On the other hand, liability to a few psychiatric traits/disorders, for example ADHD, alcohol and opioid use disorders may be causally associated with higher risks of COVID-19 infection or severe disease. In genetic correlation analysis, cannabis use disorder, ADHD, and anxiety showed significant and positive genetic correlation with critical or hospitalized infection. All the above findings passed multiple testing correction at a false discovery rate (FDR)<0.05. For pneumonia, in general we observed a different pattern of causal associations. We observed bi-directional positive associations with depression- and anxiety-related phenotypes.

**Conclusions:** In summary, this study provides evidence for tentative bi-directional causal associations between liability to COVID-19 (and severe infection) and a number of neuropsychiatric disorders. Further replications and prospective studies are required to verify the findings.

## Introduction

The number of confirmed COVID-19 has exceeded 180 million and more than 3.9 million fatalities were reported at 25 Jun 2021. A number of risk factors have been identified for COVID-19 infection and severity of infection, mostly related to age, sex and cardiometabolic disorders or abnormalities (e.g. obesity, diabetes mellitus [DM], chronic renal disease etc.). However, there has been relatively limited evidence on whether psychiatric disorders may affect the risk or severity of COVID-19.

A recent study using electronic health records in US showed that patients with schizophrenia and depression had elevated risks for infection, and as a whole, patients with mental disorders showed higher hospitalization and mortality rates^1^. In another study in Korea^2^, history of psychiatric disorders did not significantly affect the infection risk but was associated with a modest increase in the risk of severe disease (adjusted odds ratio (OR) of 1.27). Yet another recent study^3^ on mortality reported that schizophrenia spectrum disorders were associated with higher mortality, but not mood or anxiety disorders. The main limitation is that confounding may create spurious associations and render causal inference difficult. For example, psychiatric disorders are frequently associated with (lower) socioeconomic status, mental/physical comorbidities and use of various medications, all of which may also be associated with infection risk or severity.

On the other hand, it is possible that neuropsychiatric disorders may develop as a consequence of the infection. A recent large-scale study^4^ by Taquet et al. reported that COVID-19 infection is associated with higher incidence of a psychiatric diagnosis (new or recurrent) 14-90 days after a COVID-19 diagnosis. Conversely, history of psychiatric disorder within one year was associated with approximately 65% increased risk of COVID-19. Also, a study from China^5^ revealed that numerous symptoms may persist after discharge; the most common symptoms included fatigue/muscle weakness, sleep problems and anxiety or depressive symptoms. Of note, neuropsychiatric sequelae are considered an important component of the ‘long-COVID’ syndrome^6^, although studies on such consequences are still limited. Again a major limitation of these studies is that many factors may influence both the risk of infection and psychiatric disorders, leading to confounding. Also for some patients, it is possible that the psychiatric disorder remain undiagnosed until after more detailed follow-up post-infection, as a result reverse causality may also explain the association.

Here we employed Mendelian randomization (MR) to evaluate causal relationships between neuropsychiatric disorders and COVID-19 infection, including hospitalized and critical cases as subtypes. MR is much less prone to confounding and reverse causality when compared to observational studies. In addition, some of the psychiatric disorders may have relatively low incidence (e.g. schizophrenia) and may remain undetected with modest sample sizes and limited duration of follow-up. On the other hand, MR only requires summary statistics from genome-wide association studies (GWAS), which were usually of very large sample sizes (often >100,000). This could improve the statistical power to detect causal relationships of COVID-19 with neuropsychiatric disorders. Of note, prior to the outbreak of COVID-19, several studies have suggested that severe mental disorders such as schizophrenia may increase the risk of pneumonia^7–9^. We therefore also include pneumonia as an exposure and outcome in this study, as pneumonia itself is also a major public health burden and the comparison with COVID-19 will be of interest.

## Methods

### GWAS data

#### COVID-19 data

We extracted GWAS summary statistics from the COVID-19 Host Genetics Initiative, release 5 (updated Jan 18 2021). Please also refer to ref^10^, ref^11^, and https://docs.google.com/document/d/16ethjgi4MzlQeO0KAW_yDYyUHdB9kKbtfuGW4XYVKQg/edit for details on samples and analytic methodologies.

We focused on three sets of GWAS results, including very severe/critically ill, hospitalized, and (any) COVID-19 cases compared against unscreened population controls (denoted as A2, B2, C2 respectively). Detailed definitions of these outcomes are given at https://docs.google.com/document/d/1okamrqYmJfa35ClLvCt_vEe4PkvrTwggHq7T3jbeyCI/edit. Very severe or critically ill cases are defined as hospitalized and laboratory-confirmed cases who required respiratory support or whose mortality was related to the infection. These three datasets were chosen mainly because the sample sizes were among the largest (A2: 5870 cases/1,155,203 controls; B2, 11,829 cases/1,725,210 controls; C2: 42,557 cases/1,424,707 controls). We selected the sets of GWAS summary statistics that did not contain the UK Biobank (UKBB) sample, to minimize the chance of sample overlap with GWAS of neuropsychiatric disorders.

#### Pneumonia

For pneumonia, we conducted a meta-analysis on the GWAS results from the UKBB (phecode 480) and Finngen (under the code J10_pneumonia). The combined results from meta-analysis were used as input for further MR analysis.

#### Neuropsychiatric disorders

The list of neuropsychiatric disorders or traits under study is presented in Table 1. For details, please refer to the references therein. In brief, we included a wide range of psychiatric disorders or traits which includes schizophrenia (SCZ), bipolar disorder (BPD) (including bipolar disorder I, II and combined), major depressive disorder (MDD) [including ordinary MDD^12^, severe MDD requiring electroconvulsive therapy and melancholic depression from MDD-CONVERGE], other depression-related phenotypes (general depressive symptoms, number of depressive episodes, suicidal thoughts [ST], self-harm, suicide attempts[SA], insomnia), psychotic experience (PE), anxiety disorders (based on meta-analysis of PGC and UKBB samples conducted by us), general anxious symptoms (UKBB data field 1980), neuroticism (two studies), anorexia nervosa (AN), obsessive compulsive disorder (OCD), post-traumatic stress disorder (PTSD), attention deficit hyperactivity disorder (ADHD) and substance-related disorders (alcohol dependence[ALD], cannabis use disorder[CUD], and opioid dependence[OD]). For OD, for comprehensiveness, we included several comparisons including OD vs subjects exposed or unexposed to opioids, and comparison of exposed vs unexposed individuals. We also included several neurological disorders including Alzheimer’s disease, Lewy Body dementia and Parkinson’s disease.

**Table 1.** Description of GWAS data of neuropsychiatric disorders/traits used in this study.

| Phenotype | abbr. | Source | data type | Cases | Controls | Total N | No. of SNPs | PMID/Links |
| --- | --- | --- | --- | --- | --- | --- | --- | --- |
| Alcohol dependence 2018 | alco2018 | PGC | binary | 11,476 | 23,080 | 34,556 | 7,939,164 | PMID: 30482948 |
| Alzheimer disease | ad | CTG lab | binary | 71,880 | 383,378 | 455,258 | 13,367,299 | PMID: 30617256 |
| Anxiety disorders (UKBB phcode 300) | ukbb.anxiety | UKBB | binary | 10,751 | 384,235 | 394,986 | 24,385,924 | Pan-UKB |
| Anxiety disorder case control study (PGC) | pgc.anx.cc | PGC | binary | 7,016 | 14,745 | 17,310 | 6,330,995 | PMID: 26754954 |
| Anxiety disorder Meta (PGC anxiety + ukbb.anxiety) | meta.anxiety | This study | binary | 17,767 | 398,980 | 416,747 | 5,977,203 | NA |
| Worrier / anxious feelings (UKBB data field 1980) | anx | UKBB | binary | 213,808 | 162,603 | 376,411 | 10,579,925 | PMID: 31427789 |
| Attention Deficit Hyperactivity Disorder 2019 | adhd2019 | IPSYCH | binary | 19,099 | 34,194 | 53,293 | 8,047,420 | PMID: 30478444 |
| Bipolar Disorder I PGC3 2021 | bip.2021.i | PGC | binary | 250,606 | 307,499 | 558,105 | 7,391,594 | PMID: 34002096 |
| Bipolar Disorder II PGC3 2021 | bip.2021.ii | PGC | binary | 6,781 | 273,693 | 280,474 | 7,188,236 | PMID: 34002096 |
| Bipolar Disorder PGC3 All | bip.2021.all | PGC | binary | 41,917 | 371,549 | 413,466 | 7,608,183 | PMID: 34002096 |
| Cannabis Use Disorder 2020 Eur (Exclude related subjects) | cud.eur.exclude | PGC | binary | 14,808 | 343,726 | 358,534 | 8,851,634 | PMID: 33096046 |
| Depressive Symptoms | dep.sym2016 | SSGAC | continuous | --- | --- | 161,460 | 6,524,474 | PMID: 27089181 |
| Eating Disorder (anorexia nervosa) 2019 | ed2019 | PGC | binary | 16,992 | 55,525 | 72,517 | 8,219,102 | PMID: 31308545 |
| Ever contemplated self-harm (UKBB data field 20485) | ever.sh | UKBB | binary | 5,099 | 112,634 | 117,733 | 12,075,153 | Pan-UKB |
| Insomnia 2019 | insom2019 | UKBB&CTG | binary | 109,389 | 277,144 | 386,533 | 10,862,567 | PMID: 30804565 |
| Lewy Body Dementia | dementia.lbd2020 | Chia et al. | binary | 2,591 | 4,027 | 6,618 | 7,843,595 | PMID: 33589841 |
| Major Depressive Disorder 2019 | mdd2019 | PGC | binary | 170,756 | 329,443 | 500,199 | 8,483,301 | PMID: 30718901 |
| Major Depressive Disorder (severe MDD requiring ECT) 2021 Broad | mdd.2021.broad | PGC | binary | 2,725 | 3,290 | 6,015 | 7,751,438 | PMID: 33483693 |
| Major Depressive Disorder (severe MDD requiring ECT) 2021 Narrow | mdd.2021.narrow | PGC | binary | 1,796 | 3,290 | 5,086 | 7,750,957 | PMID: 33483693 |
| Major Depressive Disorder CONVERGE | mddco | PGC | binary | 5,303 | 5,337 | 10,640 | 5,992,772 | PMID: 26176920 |
| Neuroticism (SSGAC) | neuroti.meta2016 | SSGAC | continuous | --- | --- | 170,991 | 6,524,432 | PMID: 27089181 |
| Neuroticism (UKBB 2018) | neuroti_eur_ukbb_2018 | UKBB | continuous | --- | --- | 293,006 | 13,584,548 | PMID: 29942085 |
| Number of depression episodes (UKBB phcode 4260) | no.dep | UKBB | continuous | --- | --- | 45,695 | 13,356,624 | Pan-UKB |
| Obsessive-Compulsive Disorder 2018 | ocd2018 | PGC | binary | 2,688 | 7,037 | 9,725 | 8,409,516 | PMID: 28761083 |
| Opioid Dependence 2020 Case/Exposed Eur | od.case-exposed.eur | PGC | binary | 3,272 | 2,876 | 6,148 | 4,213,977 | PMID: 32099098 |
| Opioid Dependence 2020 Case/Exposed Trans | od.case-exposed.trans | PGC | binary | 4,503 | 4,173 | 8,676 | 4,002,503 | PMID: 32099098 |
| Opioid Dependence 2020 Case/Unexposed Eur | od.case-unexposed.eur | PGC | binary | 2,712 | 10,540 | 13,252 | 4,889,007 | PMID: 32099098 |
| Opioid Dependence 2020 Case/Unexposed Trans | od.case-unexposed.trans | PGC | binary | 3,594 | 15,895 | 19,489 | 4,012,235 | PMID: 32099098 |
| Opioid Dependence 2020 Exposed/Unexposed Eur | od.exposed-unexposed.eur | PGC | binary | 2,876 | 25,022 | 27,898 | 5,989,497 | PMID: 32099098 |
| Opioid Dependence 2020 Exposed/Unexposed Trans | od.exposed-unexposed.trans | PGC | binary | 4,173 | 31,820 | 35,993 | 7,435,111 | PMID: 32099098 |
| Parkinson Disease Meta5 (excluding 23andMe) | pd | IPDGC | binary | 5,542 | 5,866 | 11,408 | 17,669,774 | PMID: 25444595 |
| Pneumonia UKBB | ukbb.pneumonia | UKBB | binary | 14,507 | 423,632 | 438,139 | 25,852,522 | Pan-UKB |
| Pneumonia Meta-analysis (used in our final analysis) | meta.pneumonia | This study | binary | 70,115 | 97,305 | 573,777 | 10,996,219 | NA |
| Pneumonia FinnGen r3 J10 | finn.pneumonia | FinnGen | binary | 15,771 | 119,867 | 135,638 | 16,948,862 | FinnGen |
| Posttraumatic Stress Disorder Freeze 2 2019 | ptsd2019 | PGC | binary | 32,428 | 174,227 | 206,655 | 8,659,506 | PMID: 31594949 |
| Psychotic Experiences 2019 | psycho.experi | UKBB | binary | 6,123 | 121,843 | 127,966 | 7,562,334 | PMID: 31553412 |
| Recent thoughts of suicide or self-harm (UKBB data field 20513) | st | UKBB | continuous | --- | --- | 117,177 | 13,571,470 | Pan-UKB |
| Schizophrenia Wave3 Public.v2 | scz.wave3.v2 | PGC | binary | 67,390 | 94,015 | 161,405 | 7,481,682 | Ripke, Stephan, et al. |
| Suicide Attempts with or without Mental Disorders | sa.ipsych | IPSYCH | binary | 6,024 | 44,240 | 50,264 | 8,017,026 | PMID: 30116032 |
| Thoughts of death during worst depression (UKBB data field 20437) | st.dep | UKBB | binary | 32,630 | 30,018 | 62,648 | 13,559,507 | Pan-UKB |

UKBB, UK Biobank; MDD, major depressive disorder; ECT, electroconvulsive therapy; Eur, European; Trans, trans-ethnic. For opioid dependence, case represents subjects with opioid dependence, exposed represents those exposed to opioids.

The majority of the samples are European in ancestry, but we also included meta-analysis of trans-ethnic samples for larger sample sizes. All GWAS summary statistics were corrected for population stratification.

### Mendelian randomization analysis

Here we performed two-sample MR, in which the instrument-exposure and instrument-outcome associations were estimated in different samples.

We conducted MR with several different MR approaches, including (1) inverse-variance weighted (MR-IVW)^13^ method; (2) Egger regression (MR-Egger)^14^; (3) weighted median (WM)^15^; (4) GSMR^16^; and (5) MR-RAPS^17^. We employed the “TwoSampleMR” R package for methods (1) to (3) and the GSMR R package for method (4). For method (5), the “mr-raps” package was used with default settings, allowing for overdispersion and shrinkage estimates. Briefly, MR-IVW is a widely used and standard approach for MR, based on meta-analysis of single-SNP MR results. The MR-Egger approach allows for imbalanced horizontal pleiotropy, and all the instruments can be invalid. The statistical power is however weaker. The weighted median method employed a median estimate which allows at most half of the instruments to be invalid. GSMR employed an ‘outlier-removal’ principle to exclude SNPs that are likely invalid instruments, and is similar to MR-IVW in principle with slight differences. The MR-RAPs method employed another approach that considers the measurement error in SNP-exposure effects and has been shown to be unbiased in the presence of many weak instruments. MR-RAPS allows both systematic and idiosyncratic pleiotropy.

Each of the above methods is based on different assumptions, and the statistical power also differs. However, it is hard to evaluate a priori which MR approach is the most optimal for a certain analysis. Hence we performed analyses with multiple methods; results supported by multiple MR approaches are considered to be relatively more robust. The false discovery rate (FDR) approach was used to correct for multiple testing.

One of the major concerns of MR is (imbalanced) horizontal pleiotropy, in which the genetic instruments have effects on the target phenotype through pathways not passing through the exposure. Except MR-IVW, the other four methods can account for such pleiotropy, given that corresponding assumptions are satisfied.

#### Inclusion of a larger number of SNPs as instruments

We note that the number of SNP instruments passing genome-wide significance (*p*<5e-8) is generally small (particularly for COVID-19 as exposure), which may limit the power to detect possible causal relationships. One approach is to increase the number of instruments by relaxing the p-value threshold. This will lead to weak instrument bias, but the direction is towards the null for two-sample MR (i.e. conservative bias). A previous simulation study^18^ showed that that type I error control (for null effect) is maintained for very weak instruments^18^. Another study^19^ showed that type I error is controlled at the nominal rate when MR is conducted with up to ∼900 SNPs which fell short of genome-wide significance. We have also conducted extensive simulations earlier^20^ and have showed that relaxation of the p-value threshold (pthres) up to 0.01 does not lead to increased rate of false positive findings. Based on the above studies and recent advances, here we also considered more liberal pthres for instrument inclusion to improve power. To avoid the arbitrariness of setting particular thresholds, we considered a range of thresholds (p=5e-8, 1e-7, 1e-6, 1e-5, 1e-4, 1e-3, 1e-2) and corrected for multiple comparisons by FDR.

The other approach is to include SNPs in LD (correlated SNPs) ^21^ for MR analysis. The methodology to account for LD has been developed ^21^. However, if the SNPs are too highly correlated, the resulting estimates may be unstable^21^. Simulation studies^21^ showed that type I error was controlled at correct levels, for up to ∼320 correlated SNPs (correlation ∼0.4-0.6). To avoid unstable estimates, based on the above findings, we set an r^2^ threshold of 0.2 and a threshold for number of SNPs at 350, for analysis involving correlated SNPs. Since there is no consensus for a particular r^2^ cut-off for optimal results, we performed MR analysis with correlated SNPs at four levels of r^2^ (0.05, 0.1, 0.15 and 0.2) and assessed the consistency of results. Multiple testing was accounted for by FDR^21^. The R packages “MendelianRandomization” and “gmsr” were used for MR-IVW and GSMR of correlated SNPs respectively.

#### Interpretation of MR causal estimate for binary exposure

For exposures that are binary, the MR estimate is equal to the change in the outcome per log-odds change of the exposure. The causal estimate reflects the change of outcome for every 2.72-fold increase in the odds of the exposure. For uncommon outcomes, the MR estimate is approximately equal to 2.72-fold increase in the probability of exposure, e.g. a change of disease/infection risk from 1% to 2.72%. ^22^.

#### Steiger test of directionality

In brief, this test examines whether the instrument SNPs explain more variance for exposure than for the outcome ^23^. This serves to further confirm whether the causal direction is correct. The test is applicable to independent SNPs. We employed the mr_steiger function in “TwoSampleMR” for this test. We would primarily focus on results whose causal direction was indicated as “TRUE” by the function.

#### Multiple testing control by FDR

Multiple testing was controlled by the FDR approach, which controls the expected *proportion* of false positives among the rejected hypotheses.

In this study we set a FDR threshold of 0.05 to declare significance. Results with FDR<0.1 but >0.05 are considered ‘suggestive’ associations. FDR calculation was stratified by each psychiatric trait, which also enables average FDR (averaged across all subgroups) to be controlled^24^.

#### Genetic correlation analysis

Genetic correlation analysis was performed using LD score regression ^25^ following default settings. The method evaluates genetic overlap between pairs of disorders using the entire GWAS panel of SNPs, although it is not designed for inferring causality.

## Results

We will mainly present results that survive multiple testing correction at FDR<0.05 (considered as ‘significant’ associations in this study). Results are presented in Tables 2-9. Results of suggestive associations (0.05<FDR<0.1) are shown in Tables S1-S8 and Figures 1-2.

**Table 2.** MR results (FDR<0.05) with liability to COVID-19 as exposure and neuropsychiatric disorders as outcome (independent SNPs as instruments)

| exp | out | method | nsnps | b | se | OR | LCI | UCI | pval | p_thres | p.adjust |
| --- | --- | --- | --- | --- | --- | --- | --- | --- | --- | --- | --- |
| B2 | adhd2019 | GSMR | 136 | 0.047 | 0.014 | 1.048 | 1.019 | 1.079 | 1.10E-03 | 1.00E-04 | 3.26E-02 |
| B2 | adhd2019 | IVW | 136 | 0.051 | 0.015 | 1.052 | 1.022 | 1.083 | 5.58E-04 | 1.00E-04 | 3.26E-02 |
| B2 | adhd2019 | MR-RAPS | 136 | 0.056 | 0.017 | 1.058 | 1.024 | 1.093 | 7.02E-04 | 1.00E-04 | 3.26E-02 |
| A2 | adhd2019 | SIMEX | 401 | 0.025 | 0.007 | 1.025 | 1.011 | 1.039 | 6.83E-04 | 1.00E-03 | 3.26E-02 |
| B2 | adhd2019 | SIMEX | 136 | 0.053 | 0.016 | 1.054 | 1.022 | 1.087 | 1.05E-03 | 1.00E-04 | 3.26E-02 |
| B2 | adhd2019 | Wt-median | 136 | 0.069 | 0.021 | 1.071 | 1.028 | 1.116 | 1.12E-03 | 1.00E-04 | 3.26E-02 |
| A2 | bip.2021.ii | GSMR | 30 | 0.071 | 0.027 | 1.074 | 1.019 | 1.131 | 7.67E-03 | 1.00E-05 | 4.88E-02 |
| B2 | bip.2021.ii | GSMR | 840 | 0.036 | 0.011 | 1.036 | 1.014 | 1.060 | 1.50E-03 | 1.00E-02 | 1.69E-02 |
| B2 | bip.2021.ii | IVW | 410 | 0.040 | 0.014 | 1.041 | 1.013 | 1.070 | 3.50E-03 | 1.00E-03 | 2.75E-02 |
| B2 | bip.2021.ii | MR Egger | 840 | 0.123 | 0.032 | 1.131 | 1.063 | 1.204 | 1.15E-04 | 1.00E-02 | 5.59E-03 |
| B2 | bip.2021.ii | MR-RAPS | 410 | 0.044 | 0.015 | 1.045 | 1.014 | 1.077 | 3.82E-03 | 1.00E-03 | 2.80E-02 |
| A2 | bip.2021.ii | SIMEX | 30 | 0.084 | 0.028 | 1.088 | 1.031 | 1.148 | 4.91E-03 | 1.00E-05 | 3.24E-02 |
| A2 | ptsd2019 | IVW | 423 | 0.020 | 0.006 | 1.020 | 1.009 | 1.031 | 4.51E-04 | 1.00E-03 | 3.07E-02 |
| A2 | ptsd2019 | SIMEX | 423 | 0.023 | 0.006 | 1.024 | 1.011 | 1.036 | 1.71E-04 | 1.00E-03 | 2.85E-02 |
| B2 | scz.wave3.v2 | GSMR | 841 | 0.015 | 0.004 | 1.016 | 1.007 | 1.024 | 4.71E-04 | 1.00E-02 | 2.29E-02 |
| A2 | scz.wave3.v2 | IVW | 12 | 0.048 | 0.015 | 1.049 | 1.019 | 1.080 | 1.13E-03 | 1.00E-06 | 3.14E-02 |
| B2 | scz.wave3.v2 | IVW | 846 | 0.019 | 0.006 | 1.019 | 1.008 | 1.030 | 6.33E-04 | 1.00E-02 | 2.50E-02 |
| B2 | scz.wave3.v2 | MR-RAPS | 846 | 0.019 | 0.006 | 1.020 | 1.008 | 1.032 | 1.16E-03 | 1.00E-02 | 3.14E-02 |
| B2 | scz.wave3.v2 | SIMEX | 846 | 0.020 | 0.006 | 1.021 | 1.009 | 1.033 | 7.20E-04 | 1.00E-02 | 2.50E-02 |
| A2 | st | GSMR | 11 | 0.007 | 0.002 | --- | 0.003 | 0.011 | 2.85E-03 | 1.00E-06 | 4.27E-02 |
| B2 | st | GSMR | 9 | 0.014 | 0.004 | --- | 0.006 | 0.022 | 2.45E-04 | 1.00E-06 | 6.43E-03 |
| A2 | st | SIMEX | 11 | 0.009 | 0.002 | --- | 0.005 | 0.013 | 3.64E-03 | 1.00E-06 | 4.92E-02 |
| B2 | st | SIMEX | 9 | 0.015 | 0.004 | --- | 0.007 | 0.023 | 5.37E-03 | 1.00E-06 | 4.92E-02 |

For each exposure-outcome-method combination, we only show at most one result (selecting the result with lowest p\_thres and r2 threshold). Only results which passed the Steiger test of directionality are shown in main tables.
Note: od.cu.e: od.case\_unexposed.eur; od.ce.e: od.case\_exposure.eur; od.expu.e: od.case\_exposed.eur; od.expu.t: od.exposed\_unexposed.trans;

**Table 3.** MR results (FDR<0.05) with liability to COVID-19 as exposure and neuropsychiatric disorders as outcome (correlated SNPs as instruments)

| exp | out | method | nsnps | b | se | OR | LCI | UCI | pval | p_thres | r2 | p.adjust |
| --- | --- | --- | --- | --- | --- | --- | --- | --- | --- | --- | --- | --- |
| B2 | bip.2021.all | IVW | 201 | 0.042 | 0.010 | 1.043 | 1.023 | 1.063 | 1.58E-05 | 1.00E-04 | 0.05 | 9.05E-04 |
| A2 | bip.2021.ii | GSMR | 36 | 0.076 | 0.025 | 1.079 | 1.028 | 1.133 | 2.10E-03 | 1.00E-05 | 0.05 | 2.02E-02 |
| A2 | bip.2021.ii | IVW | 36 | 0.088 | 0.024 | 1.092 | 1.042 | 1.143 | 2.09E-04 | 1.00E-05 | 0.05 | 5.59E-03 |
| B2 | bip.2021.ii | IVW | 37 | 0.096 | 0.034 | 1.101 | 1.031 | 1.176 | 4.35E-03 | 1.00E-05 | 0.05 | 3.00E-02 |
| B2 | bip.2021.ii | MR Egger | 199 | 0.160 | 0.044 | 1.173 | 1.076 | 1.279 | 3.00E-04 | 1.00E-04 | 0.05 | 5.59E-03 |
| C2 | meta.anxiety | MR Egger | 156 | 0.286 | 0.074 | 1.331 | 1.151 | 1.539 | 1.16E-04 | 1.00E-04 | 0.1 | 1.14E-02 |
| B2 | ocd2018 | MR Egger | 202 | 0.237 | 0.067 | 1.267 | 1.112 | 1.444 | 3.78E-04 | 1.00E-04 | 0.05 | 4.91E-02 |
| B2 | ptsd2019 | MR Egger | 224 | 0.091 | 0.025 | 1.096 | 1.043 | 1.151 | 2.80E-04 | 1.00E-04 | 0.2 | 2.85E-02 |
| B2 | scz.wave3.v2 | IVW | 201 | 0.048 | 0.010 | 1.049 | 1.029 | 1.069 | 1.17E-06 | 1.00E-04 | 0.05 | 1.51E-04 |
| B2 | st | GSMR | 9 | 0.014 | 0.004 | --- | 0.006 | 0.022 | 2.45E-04 | 1.00E-06 | 0.05 | 6.43E-03 |
| A2 | st | MR Egger | 8 | 0.034 | 0.012 | --- | 0.010 | 0.058 | 5.01E-03 | 1.00E-07 | 0.15 | 4.92E-02 |
| B2 | st | MR Egger | 6 | 0.053 | 0.019 | --- | 0.016 | 0.090 | 5.21E-03 | 1.00E-07 | 0.05 | 4.92E-02 |

R2, LD-clumping threshold for inclusion as instruments. Note: neuroti\_ukbb: neuroti\_eur\_ukbb\_2018; od.expu.t: od.exposed\_unexposed.trans.

**Table 4.** MR results (FDR<0.05) with liability to neuropsychiatric disorders as exposure and COVID-19 as outcome (independent SNPs as instruments)

| exp | out | method | nsnps | b | se | OR | LCI | UCI | pval | p_thres | p.adjust |
| --- | --- | --- | --- | --- | --- | --- | --- | --- | --- | --- | --- |
| alco2018.beta | C2 | GSMR | 113 | 0.035 | 0.011 | 1.036 | 1.014 | 1.057 | 8.68E-04 | 1.00E-04 | 7.74E-03 |
| alco2018.beta | C2 | IVW | 113 | 0.038 | 0.010 | 1.039 | 1.019 | 1.060 | 1.66E-04 | 1.00E-04 | 4.45E-03 |
| alco2018.beta | C2 | MR-RAPS | 113 | 0.040 | 0.011 | 1.041 | 1.018 | 1.063 | 3.59E-04 | 1.00E-04 | 6.69E-03 |
| alco2018.beta | C2 | SIMEX | 113 | 0.037 | 0.011 | 1.037 | 1.015 | 1.060 | 1.28E-03 | 1.00E-04 | 1.06E-02 |
| od.cu.t | B2 | IVW | 654 | 0.001 | 0.000 | 1.001 | 1.000 | 1.002 | 3.95E-03 | 1.00E-02 | 2.16E-02 |
| od.cu.t | C2 | IVW | 227 | 0.001 | 0.000 | 1.001 | 1.000 | 1.001 | 8.54E-03 | 1.00E-03 | 3.91E-02 |
| od.cu.t | B2 | MR-RAPS | 654 | 0.003 | 0.001 | 1.003 | 1.001 | 1.006 | 3.54E-03 | 1.00E-02 | 2.16E-02 |
| od.cu.t | C2 | MR-RAPS | 227 | 0.002 | 0.001 | 1.002 | 1.001 | 1.003 | 4.39E-03 | 1.00E-03 | 2.16E-02 |
| od.cu.t | A2 | SIMEX | 652 | 0.003 | 0.001 | 1.003 | 1.001 | 1.005 | 3.39E-04 | 1.00E-02 | 5.43E-03 |
| od.cu.t | B2 | SIMEX | 68 | 0.008 | 0.002 | 1.008 | 1.003 | 1.012 | 2.77E-03 | 1.00E-04 | 2.16E-02 |
| od.cu.t | C2 | SIMEX | 15 | 0.017 | 0.005 | 1.017 | 1.008 | 1.027 | 3.39E-03 | 1.00E-05 | 2.16E-02 |
| od.expu.e | C2 | SIMEX | 196 | 0.002 | 0.001 | 1.002 | 1.001 | 1.004 | 9.96E-05 | 1.00E-03 | 9.36E-03 |
| ptsd2019 | C2 | MR Egger | 994 | 0.051 | 0.016 | 1.053 | 1.021 | 1.086 | 1.16E-03 | 1.00E-02 | 4.40E-02 |
| sa.ipsych | A2 | IVW | 326 | 0.061 | 0.019 | 1.063 | 1.025 | 1.104 | 1.18E-03 | 1.00E-03 | 3.87E-02 |
| sa.ipsych | C2 | MR Egger | 350 | 0.045 | 0.014 | 1.046 | 1.018 | 1.075 | 1.35E-03 | 1.00E-03 | 3.87E-02 |
| sa.ipsych | A2 | MR-RAPS | 326 | 0.069 | 0.021 | 1.071 | 1.028 | 1.116 | 1.01E-03 | 1.00E-03 | 3.87E-02 |
| sa.ipsych | A2 | SIMEX | 326 | 0.069 | 0.020 | 1.071 | 1.029 | 1.115 | 8.04E-04 | 1.00E-03 | 3.87E-02 |

Note: od.cu.t: od.case\_unexposed.trans; od.cu.e: od.case\_unexposed.eur; od.ce.e: od.case\_exposure.eur; od.expu.e: od.case\_exposed.eur; od.expu.t: od.exposed\_unexposed.trans; neuroti\_eur: neuroti\_eur\_ukbb\_2018;

**Table 5.** MR results (FDR<0.05) with liability to neuropsychiatric disorders as exposure and COVID-19 as outcome (correlated SNPs as instruments)

| exp | out | method | nsnps | b | se | OR | LCI | UCI | pval | p_thres | r2 | p.adjust |
| --- | --- | --- | --- | --- | --- | --- | --- | --- | --- | --- | --- | --- |
| ad | A2 | GSMR | 87 | -0.300 | 0.086 | 0.741 | 0.625 | 0.877 | 5.01E-04 | 1.00E-07 | 0.2 | 3.92E-02 |
| ad | A2 | IVW | 301 | -0.380 | 0.111 | 0.684 | 0.551 | 0.85 | 5.96E-04 | 1.00E-04 | 0.05 | 3.92E-02 |
| ad | A2 | MR Egger | 301 | -0.493 | 0.130 | 0.611 | 0.473 | 0.788 | 1.53E-04 | 1.00E-04 | 0.05 | 2.01E-02 |
| adhd2019 | A2 | IVW | 327 | 0.106 | 0.031 | 1.112 | 1.046 | 1.182 | 7.20E-04 | 1.00E-04 | 0.1 | 2.29E-02 |
| adhd2019 | A2 | MR Egger | 313 | 0.222 | 0.069 | 1.249 | 1.092 | 1.429 | 1.21E-03 | 1.00E-04 | 0.05 | 3.44E-02 |
| adhd2019 | B2 | MR Egger | 314 | 0.193 | 0.045 | 1.213 | 1.11 | 1.326 | 1.99E-05 | 1.00E-04 | 0.05 | 1.01E-03 |
| alco2018.beta | C2 | GSMR | 153 | 0.032 | 0.009 | 1.032 | 1.014 | 1.051 | 6.25E-04 | 1.00E-04 | 0.05 | 6.69E-03 |
| alco2018.beta | C2 | IVW | 153 | 0.044 | 0.009 | 1.045 | 1.027 | 1.064 | 1.06E-06 | 1.00E-04 | 0.05 | 4.30E-05 |
| alco2018.beta | A2 | MR Egger | 153 | 0.190 | 0.070 | 1.209 | 1.055 | 1.386 | 6.41E-03 | 1.00E-04 | 0.05 | 4.57E-02 |
| alco2018.beta | C2 | MR Egger | 153 | 0.085 | 0.025 | 1.089 | 1.038 | 1.143 | 5.28E-04 | 1.00E-04 | 0.05 | 6.69E-03 |
| anx | B2 | MR Egger | 259 | -0.671 | 0.194 | 0.511 | 0.349 | 0.747 | 5.31E-04 | 1.00E-05 | 0.05 | 2.49E-02 |
| anx | C2 | MR Egger | 266 | -0.378 | 0.089 | 0.685 | 0.575 | 0.816 | 2.23E-05 | 1.00E-05 | 0.05 | 1.34E-03 |
| neuroti_eur | A2 | GSMR | 83 | -0.913 | 0.200 | --- | -1.305 | -0.521 | 4.73E-06 | 5.00E-08 | 0.05 | 1.23E-04 |
| neuroti_eur | B2 | GSMR | 103 | -0.333 | 0.133 | --- | -0.594 | -0.072 | 1.26E-02 | 5.00E-08 | 0.1 | 3.58E-02 |
| neuroti_eur | C2 | GSMR | 170 | -4.159 | 0.386 | --- | -4.916 | -3.402 | 5.00E-27 | 1.00E-07 | 0.15 | 7.35E-25 |
| neuroti_eur | A2 | IVW | 83 | -0.812 | 0.212 | --- | -1.228 | -0.396 | 1.27E-04 | 5.00E-08 | 0.05 | 9.60E-04 |
| neuroti_eur | A2 | MR Egger | 159 | -1.863 | 0.555 | --- | -2.951 | -0.775 | 7.97E-04 | 5.00E-08 | 0.2 | 4.19E-03 |
| neuroti_eur | B2 | MR Egger | 85 | -1.605 | 0.580 | --- | -2.742 | -0.468 | 5.65E-03 | 5.00E-08 | 0.05 | 2.08E-02 |
| od.cu.t | B2 | IVW | 75 | 0.004 | 0.001 | 1.004 | 1.001 | 1.006 | 4.09E-03 | 1.00E-04 | 0.05 | 2.16E-02 |
| ptsd2019 | C2 | MR Egger | 248 | 0.127 | 0.025 | 1.136 | 1.081 | 1.194 | 5.33E-07 | 1.00E-04 | 0.1 | 6.08E-05 |

Note: neuroti\_eur: neuroti\_eur\_ukbb\_2018; od.cu.t: od.case-unexposed.trans

**Table 6.** MR results (FDR<0.05) with liability to pneumonia as exposure and neuropsychiatric disorders as outcome (independent SNPs as instruments)

| exp | out | method | nsnps | b | se | OR | LCI | UCI | pval | p_thres | p.adjust |
| --- | --- | --- | --- | --- | --- | --- | --- | --- | --- | --- | --- |
| FP | cud.eur.exclude | GSMR | 854 | 0.066 | 0.022 | 1.068 | 1.023 | 1.115 | 2.67E-03 | 1.00E-02 | 1.22E-02 |
| FP | cud.eur.exclude | IVW | 103 | 0.132 | 0.052 | 1.141 | 1.031 | 1.262 | 1.11E-02 | 1.00E-04 | 3.96E-02 |
| FP | cud.eur.exclude | MR Egger | 855 | 0.140 | 0.047 | 1.150 | 1.050 | 1.261 | 2.83E-03 | 1.00E-02 | 1.22E-02 |
| FP | cud.eur.exclude | MR-RAPS | 389 | 0.080 | 0.031 | 1.083 | 1.019 | 1.151 | 1.04E-02 | 1.00E-03 | 3.96E-02 |
| FP | cud.eur.exclude | SIMEX | 389 | 0.097 | 0.029 | 1.102 | 1.040 | 1.167 | 1.02E-03 | 1.00E-03 | 5.54E-03 |
| FP | dep.sym2016 | GSMR | 337 | 0.023 | 0.007 | --- | 0.009 | 0.037 | 3.02E-04 | 1.00E-03 | 2.19E-03 |
| FP | dep.sym2016 | SIMEX | 83 | 0.030 | 0.012 | --- | 0.006 | 0.054 | 1.69E-02 | 1.00E-04 | 4.47E-02 |
| FP | insom2019 | GSMR | 905 | 0.018 | 0.006 | 1.018 | 1.007 | 1.030 | 2.24E-03 | 1.00E-02 | 1.85E-02 |
| FP | insom2019 | IVW | 905 | 0.021 | 0.006 | 1.021 | 1.009 | 1.033 | 4.96E-04 | 1.00E-02 | 7.69E-03 |
| FP | insom2019 | MR-RAPS | 905 | 0.023 | 0.007 | 1.023 | 1.010 | 1.037 | 6.99E-04 | 1.00E-02 | 7.69E-03 |
| FP | insom2019 | SIMEX | 421 | 0.024 | 0.009 | 1.024 | 1.007 | 1.041 | 5.73E-03 | 1.00E-03 | 3.78E-02 |
| FP | mdd2019 | GSMR | 116 | 0.032 | 0.011 | 1.032 | 1.010 | 1.055 | 4.15E-03 | 1.00E-04 | 1.50E-02 |
| FP | mdd2019 | IVW | 116 | 0.037 | 0.012 | 1.038 | 1.014 | 1.062 | 1.62E-03 | 1.00E-04 | 7.27E-03 |
| FP | mdd2019 | MR-RAPS | 116 | 0.036 | 0.012 | 1.037 | 1.012 | 1.063 | 3.54E-03 | 1.00E-04 | 1.42E-02 |
| FP | mdd2019 | SIMEX | 116 | 0.042 | 0.013 | 1.043 | 1.017 | 1.068 | 1.15E-03 | 1.00E-04 | 5.93E-03 |
| FP | mdd2019 | Wt-median | 920 | 0.017 | 0.007 | 1.017 | 1.003 | 1.031 | 2.08E-02 | 1.00E-02 | 4.68E-02 |
| FP | meta.anxiety | GSMR | 76 | 0.107 | 0.040 | 1.113 | 1.029 | 1.204 | 7.32E-03 | 1.00E-04 | 1.97E-02 |
| FP | meta.anxiety | IVW | 310 | 0.103 | 0.024 | 1.108 | 1.057 | 1.162 | 2.14E-05 | 1.00E-03 | 1.84E-04 |
| FP | meta.anxiety | MR-RAPS | 310 | 0.110 | 0.027 | 1.117 | 1.060 | 1.177 | 3.51E-05 | 1.00E-03 | 2.16E-04 |
| FP | meta.anxiety | SIMEX | 76 | 0.129 | 0.042 | 1.138 | 1.048 | 1.235 | 2.85E-03 | 1.00E-04 | 8.76E-03 |
| FP | meta.anxiety | Wt-median | 310 | 0.101 | 0.033 | 1.106 | 1.037 | 1.180 | 2.12E-03 | 1.00E-03 | 7.02E-03 |
| FP | neuroti_eur | GSMR | 126 | 0.017 | 0.006 | --- | 0.005 | 0.029 | 7.37E-03 | 1.00E-04 | 2.35E-02 |
| FP | neuroti_eur | IVW | 126 | 0.020 | 0.007 | --- | 0.006 | 0.034 | 5.91E-03 | 1.00E-04 | 2.31E-02 |
| FP | neuroti_eur | MR Egger | 966 | 0.021 | 0.006 | --- | 0.009 | 0.033 | 8.87E-04 | 1.00E-02 | 4.63E-03 |
| FP | neuroti_eur | MR-RAPS | 126 | 0.020 | 0.007 | --- | 0.006 | 0.034 | 7.88E-03 | 1.00E-04 | 2.35E-02 |
| FP | neuroti_eur | SIMEX | 126 | 0.021 | 0.008 | --- | 0.005 | 0.037 | 8.00E-03 | 1.00E-04 | 2.35E-02 |
| FP | neuroti_eur | Wt-median | 966 | 0.013 | 0.004 | --- | 0.005 | 0.021 | 3.94E-03 | 1.00E-02 | 1.68E-02 |
| FP | neuroti.meta2016 | GSMR | 83 | 0.026 | 0.011 | --- | 0.004 | 0.048 | 1.77E-02 | 1.00E-04 | 4.70E-02 |
| FP | neuroti.meta2016 | IVW | 83 | 0.029 | 0.011 | --- | 0.007 | 0.051 | 8.03E-03 | 1.00E-04 | 2.60E-02 |
| FP | neuroti.meta2016 | MR-RAPS | 83 | 0.030 | 0.012 | --- | 0.006 | 0.054 | 1.04E-02 | 1.00E-04 | 2.92E-02 |
| FP | neuroti.meta2016 | SIMEX | 83 | 0.030 | 0.011 | --- | 0.008 | 0.052 | 7.44E-03 | 1.00E-04 | 2.60E-02 |

FP: FinnGen and UKBB meta-analysed Pneumonia GWAS data.

**Table 7.** MR results (FDR<0.05) with liability to pneumonia as exposure and neuropsychiatric disorders as outcome (correlated SNPs as instruments)

| exp | out | method | nsnps | b | se | OR | LCI | UCI | pval | p_thres | r2 | p.adjust |
| --- | --- | --- | --- | --- | --- | --- | --- | --- | --- | --- | --- | --- |
| FP | cud.eur.exclude | IVW | 129 | 0.157 | 0.045 | 1.170 | 1.071 | 1.278 | 5.12E-04 | 1.00E-04 | 0.05 | 4.40E-03 |
| FP | dep.sym2016 | GSMR | 100 | 0.032 | 0.011 | --- | 0.010 | 0.054 | 3.18E-03 | 1.00E-04 | 0.05 | 1.02E-02 |
| FP | dep.sym2016 | IVW | 4 | 0.106 | 0.038 | --- | 0.032 | 0.180 | 5.83E-03 | 1.00E-06 | 0.05 | 1.69E-02 |
| FP | mdd2019 | GSMR | 148 | 0.023 | 0.010 | 1.024 | 1.004 | 1.044 | 1.88E-02 | 1.00E-04 | 0.05 | 4.51E-02 |
| FP | mdd2019 | IVW | 17 | 0.057 | 0.024 | 1.059 | 1.009 | 1.110 | 1.88E-02 | 1.00E-05 | 0.05 | 4.51E-02 |
| FP | meta.anxiety | GSMR | 93 | 0.129 | 0.037 | 1.138 | 1.059 | 1.223 | 4.36E-04 | 1.00E-04 | 0.05 | 1.87E-03 |
| FP | meta.anxiety | IVW | 93 | 0.106 | 0.037 | 1.112 | 1.033 | 1.197 | 4.55E-03 | 1.00E-04 | 0.05 | 1.30E-02 |
| FP | neuroti.meta2016 | GSMR | 100 | 0.028 | 0.010 | --- | 0.008 | 0.048 | 6.70E-03 | 1.00E-04 | 0.05 | 2.60E-02 |
| FP | neuroti.meta2016 | IVW | 100 | 0.031 | 0.009 | --- | 0.013 | 0.049 | 1.02E-03 | 1.00E-04 | 0.05 | 6.53E-03 |

Note: FP: pneumonia.

**Table 8.** MR results (FDR<0.05) with liability to neuropsychiatric disorders as exposure and pneumonia as outcome (Independ SNPs as instruments)

| exp | out | method | nsnps | b | se | OR | LCI | UCI | pval | p_thres | p.adjust |
| --- | --- | --- | --- | --- | --- | --- | --- | --- | --- | --- | --- |
| adhd2019 | FP | GSMR | 9 | 0.107 | 0.040 | 1.113 | 1.028 | 1.204 | 8.26E-03 | 5.00E-08 | 1.26E-02 |
| adhd2019 | FP | IVW | 9 | 0.109 | 0.039 | 1.115 | 1.032 | 1.205 | 5.80E-03 | 5.00E-08 | 9.53E-03 |
| adhd2019 | FP | MR-RAPS | 9 | 0.118 | 0.042 | 1.125 | 1.036 | 1.221 | 5.02E-03 | 5.00E-08 | 8.74E-03 |
| adhd2019 | FP | SIMEX | 9 | 0.111 | 0.038 | 1.118 | 1.037 | 1.204 | 2.22E-02 | 5.00E-08 | 3.08E-02 |
| adhd2019 | FP | Wt-median | 9 | 0.140 | 0.052 | 1.150 | 1.040 | 1.273 | 6.52E-03 | 5.00E-08 | 1.05E-02 |
| bip.2021.i | FP | GSMR | 37 | -0.051 | 0.021 | 0.950 | 0.913 | 0.990 | 1.34E-02 | 5.00E-08 | 3.18E-02 |
| bip.2021.i | FP | IVW | 37 | -0.053 | 0.022 | 0.948 | 0.908 | 0.990 | 1.49E-02 | 5.00E-08 | 3.44E-02 |
| bip.2021.i | FP | MR-RAPS | 37 | -0.054 | 0.022 | 0.947 | 0.907 | 0.990 | 1.58E-02 | 5.00E-08 | 3.54E-02 |
| bip.2021.i | FP | SIMEX | 37 | -0.059 | 0.024 | 0.943 | 0.900 | 0.988 | 1.90E-02 | 5.00E-08 | 3.86E-02 |
| bip.2021.i | FP | Wt-median | 37 | -0.074 | 0.029 | 0.928 | 0.877 | 0.983 | 1.14E-02 | 5.00E-08 | 3.00E-02 |
| cud.eur.exclude | FP | GSMR | 6 | 0.133 | 0.048 | 1.143 | 1.040 | 1.256 | 5.57E-03 | 1.00E-07 | 1.75E-02 |
| cud.eur.exclude | FP | IVW | 2 | 0.237 | 0.083 | 1.268 | 1.077 | 1.493 | 4.35E-03 | 5.00E-08 | 1.75E-02 |
| cud.eur.exclude | FP | MR-RAPS | 2 | 0.239 | 0.093 | 1.270 | 1.059 | 1.522 | 9.91E-03 | 5.00E-08 | 2.66E-02 |
| cud.eur.exclude | FP | SIMEX | 6 | 0.135 | 0.041 | 1.145 | 1.057 | 1.240 | 2.98E-02 | 1.00E-07 | 4.68E-02 |
| cud.eur.exclude | FP | Wt-median | 6 | 0.130 | 0.060 | 1.139 | 1.013 | 1.281 | 2.95E-02 | 1.00E-07 | 4.68E-02 |
| dep.sym2016 | FP | GSMR | 13 | 0.403 | 0.147 | --- | 0.115 | 0.691 | 6.33E-03 | 1.00E-06 | 3.51E-02 |
| dep.sym2016 | FP | IVW | 13 | 0.443 | 0.143 | --- | 0.163 | 0.723 | 1.92E-03 | 1.00E-06 | 2.35E-02 |
| dep.sym2016 | FP | MR-RAPS | 13 | 0.447 | 0.157 | --- | 0.139 | 0.755 | 4.38E-03 | 1.00E-06 | 3.20E-02 |
| dep.sym2016 | FP | SIMEX | 40 | 0.313 | 0.112 | --- | 0.093 | 0.533 | 8.23E-03 | 1.00E-05 | 4.18E-02 |
| insom2019 | FP | GSMR | 566 | 0.080 | 0.020 | 1.084 | 1.042 | 1.126 | 4.89E-05 | 1.00E-03 | 4.59E-04 |
| insom2019 | FP | IVW | 273 | 0.076 | 0.025 | 1.078 | 1.027 | 1.133 | 2.57E-03 | 1.00E-04 | 1.75E-02 |
| insom2019 | FP | MR Egger | 1000 | 0.100 | 0.038 | 1.105 | 1.025 | 1.190 | 8.98E-03 | 1.00E-02 | 4.81E-02 |
| insom2019 | FP | MR-RAPS | 273 | 0.075 | 0.028 | 1.078 | 1.021 | 1.139 | 6.77E-03 | 1.00E-04 | 3.91E-02 |
| insom2019 | FP | SIMEX | 273 | 0.082 | 0.026 | 1.086 | 1.033 | 1.142 | 1.48E-03 | 1.00E-04 | 1.11E-02 |
| insom2019 | FP | Wt-median | 566 | 0.079 | 0.028 | 1.082 | 1.024 | 1.143 | 4.96E-03 | 1.00E-03 | 3.10E-02 |
| mdd2019 | FP | GSMR | 43 | 0.124 | 0.056 | 1.132 | 1.013 | 1.264 | 2.80E-02 | 1.00E-07 | 4.89E-02 |
| mdd2019 | FP | IVW | 43 | 0.131 | 0.056 | 1.140 | 1.022 | 1.271 | 1.86E-02 | 1.00E-07 | 3.66E-02 |
| mdd2019 | FP | MR-RAPS | 43 | 0.138 | 0.059 | 1.148 | 1.022 | 1.290 | 2.00E-02 | 1.00E-07 | 3.85E-02 |
| mdd2019 | FP | SIMEX | 91 | 0.166 | 0.045 | 1.181 | 1.082 | 1.289 | 3.62E-04 | 1.00E-06 | 1.11E-03 |
| mdd2019 | FP | Wt-median | 346 | 0.094 | 0.037 | 1.099 | 1.021 | 1.182 | 1.14E-02 | 1.00E-04 | 2.45E-02 |
| meta.anxiety | FP | IVW | 114 | 0.046 | 0.016 | 1.047 | 1.014 | 1.080 | 4.26E-03 | 1.00E-04 | 1.41E-02 |
| meta.anxiety | FP | MR-RAPS | 114 | 0.050 | 0.017 | 1.051 | 1.016 | 1.087 | 4.22E-03 | 1.00E-04 | 1.41E-02 |
| meta.anxiety | FP | Wt-median | 395 | 0.038 | 0.014 | 1.039 | 1.010 | 1.069 | 7.99E-03 | 1.00E-03 | 2.40E-02 |
| neuroti_eur | FP | GSMR | 274 | 0.101 | 0.043 | --- | 0.017 | 0.185 | 1.83E-02 | 1.00E-05 | 4.51E-02 |
| neuroti_eur | FP | IVW | 274 | 0.112 | 0.042 | --- | 0.030 | 0.194 | 7.83E-03 | 1.00E-05 | 2.16E-02 |
| neuroti_eur | FP | MR-RAPS | 470 | 0.151 | 0.039 | --- | 0.075 | 0.227 | 8.64E-05 | 1.00E-04 | 4.97E-04 |
| neuroti_eur | FP | SIMEX | 274 | 0.121 | 0.043 | --- | 0.037 | 0.205 | 5.36E-03 | 1.00E-05 | 1.61E-02 |
| od.case_unexposed.eur | FP | IVW | 190 | 0.001 | 0.000 | 1.001 | 1.000 | 1.001 | 1.65E-03 | 1.00E-03 | 3.96E-02 |

Note: FP: pneumonia.

**Table 9.** MR results (FDR<0.05) with liability to neuropsychiatric disorders as exposure and pneumonia as outcome (correlated SNPs as instruments)

| exp | out | method | nsnps | b | se | OR | LCI | UCI | pval | p_thres | r2 | p.adjust |
| --- | --- | --- | --- | --- | --- | --- | --- | --- | --- | --- | --- | --- |
| ad | FP | GSMR | 104 | 1.134 | 0.063 | 3.109 | 2.746 | 3.521 | 2.01E-71 | 1.00E-07 | 0.2 | 1.75E-69 |
| adhd2019 | FP | GSMR | 10 | 0.091 | 0.039 | 1.095 | 1.015 | 1.181 | 1.87E-02 | 5.00E-08 | 0.05 | 2.67E-02 |
| adhd2019 | FP | IVW | 10 | 0.093 | 0.038 | 1.097 | 1.017 | 1.183 | 1.59E-02 | 5.00E-08 | 0.05 | 2.39E-02 |
| bip.2021.i | FP | GSMR | 41 | -0.056 | 0.020 | 0.946 | 0.909 | 0.984 | 5.32E-03 | 5.00E-08 | 0.05 | 2.13E-02 |
| bip.2021.i | FP | IVW | 41 | -0.052 | 0.019 | 0.949 | 0.913 | 0.986 | 6.95E-03 | 5.00E-08 | 0.05 | 2.40E-02 |
| bip.2021.i | FP | MR Egger | 119 | -0.150 | 0.063 | 0.861 | 0.760 | 0.974 | 1.74E-02 | 1.00E-06 | 0.15 | 3.75E-02 |
| cud.eur.exclude | FP | GSMR | 6 | 0.133 | 0.048 | 1.143 | 1.040 | 1.256 | 5.57E-03 | 1.00E-07 | 0.05 | 1.75E-02 |
| cud.eur.exclude | FP | IVW | 2 | 0.237 | 0.084 | 1.267 | 1.076 | 1.493 | 4.61E-03 | 5.00E-08 | 0.05 | 1.75E-02 |
| dep.sym2016 | FP | IVW | 14 | 0.421 | 0.144 | --- | 0.139 | 0.703 | 3.45E-03 | 1.00E-06 | 0.05 | 3.20E-02 |
| mdd2019 | FP | GSMR | 54 | 0.123 | 0.052 | 1.131 | 1.022 | 1.252 | 1.73E-02 | 1.00E-07 | 0.05 | 3.50E-02 |
| mdd2019 | FP | IVW | 54 | 0.109 | 0.049 | 1.115 | 1.012 | 1.228 | 2.75E-02 | 1.00E-07 | 0.05 | 4.89E-02 |
| mdd2019 | FP | MR Egger | 121 | -0.354 | 0.138 | 0.702 | 0.535 | 0.921 | 1.05E-02 | 1.00E-06 | 0.05 | 2.38E-02 |
| meta.anxiety | FP | GSMR | 160 | 0.046 | 0.014 | 1.047 | 1.019 | 1.077 | 1.07E-03 | 1.00E-04 | 0.05 | 7.04E-03 |
| meta.anxiety | FP | IVW | 160 | 0.038 | 0.015 | 1.039 | 1.009 | 1.070 | 1.13E-02 | 1.00E-04 | 0.05 | 2.86E-02 |
| neuroti_eur_ukbb_2018 | FP | GSMR | 145 | 0.139 | 0.053 | --- | 0.035 | 0.243 | 8.35E-03 | 1.00E-07 | 0.1 | 2.22E-02 |
| neuroti_eur_ukbb_2018 | FP | IVW | 91 | 0.193 | 0.068 | --- | 0.060 | 0.326 | 4.51E-03 | 5.00E-08 | 0.05 | 1.41E-02 |
| scz.wave3.v2 | FP | MR Egger | 251 | 0.117 | 0.037 | 1.124 | 1.045 | 1.208 | 1.60E-03 | 5.00E-08 | 0.05 | 4.57E-02 |
| st | FP | GSMR | 196 | 0.308 | 0.126 | --- | 0.061 | 0.555 | 1.47E-02 | 1.00E-04 | 0.05 | 3.35E-02 |
| st | FP | IVW | 196 | 0.363 | 0.118 | --- | 0.132 | 0.594 | 2.03E-03 | 1.00E-04 | 0.05 | 1.93E-02 |

Note: FP: pneumonia.

**Figure 1.**
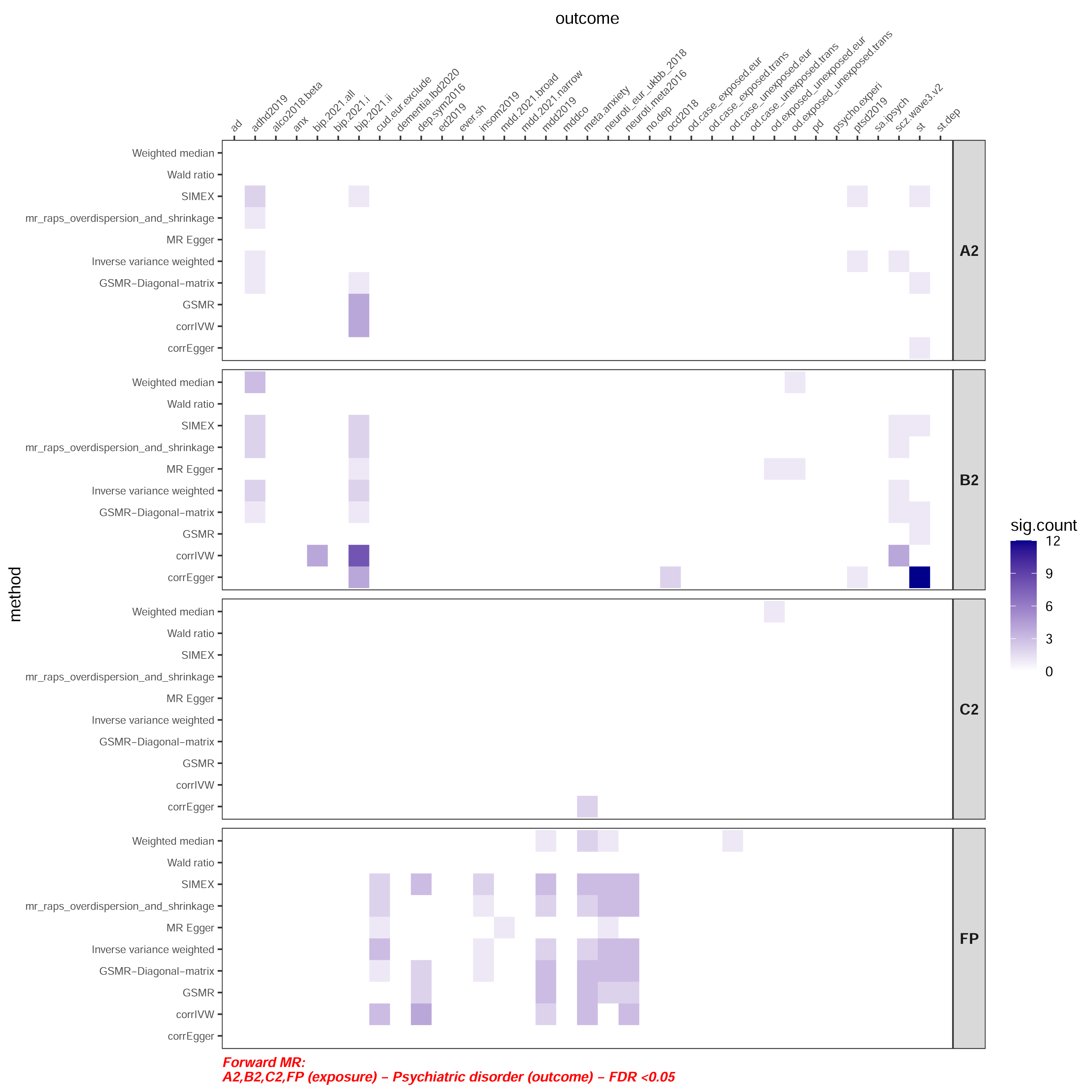
An overview of significant MR results (FDR<0.05) with COVID-19/pneumonia as exposure and neuropsychiatric disorders/traits as outcome. The color indicates the number of significant results for the specific exposure and outcome.

**Figure 2.**
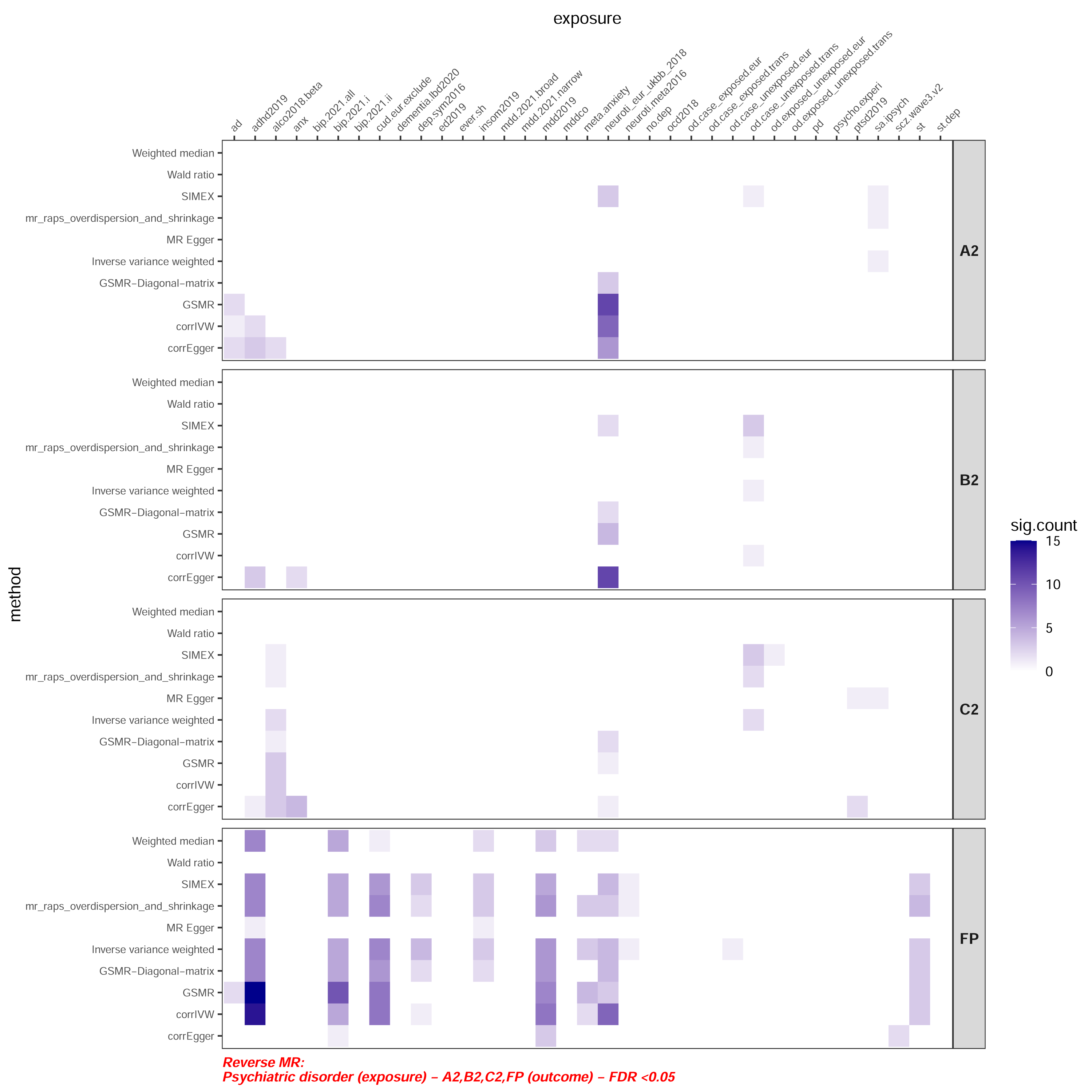
An overview of significant MR results (FDR<0.05) with neuropsychiatric disorders/traits as exposure and COVID-19/pneumonia as outcome. The color indicates the number of significant results for the specific exposure and outcome.

If significant results are observed across multiple p-value thresholds and/or r^2^ thresholds, we would present findings corresponding to the smallest p-value threshold and lowest r^2^ in the main tables. For space limits, we mainly present MR-IVW results as it is one of the most widely used approaches; full results can be found in main and supplementary tables.

### MR with liability to COVID-19 as exposure and neuropsychiatric disorders (psyD) as outcome

#### Independent SNPs (r^2^=0.001) as instruments

In the analysis with independent SNPs (*r*^2^=0.001) as exposure, liability to COVID-19 and hospitalized/critical infection was found to be causally and positively associated with several psyD. At an FDR threshold of 0.05, most significant associations were observed when A2 (critical disease) or B2 (hospitalized cases) were considered as the exposure.

All the associations were in the positive direction (COVID-19 increasing the risks of psyD). The most consistent associations were observed for ADHD and Bipolar Disorder II (associations observed across multiple MR methods and pthres), but we also observed associations with other psychiatric traits/disorders including SCZ, PTSD and suicidal thoughts (ST). Associations with the above traits were more consistent at an FDR threshold of 0.1 (Table S1).

For ADHD, significant associations were observed across all MR methods and multiple pthres. With critical illness (A2) as exposure, the OR was 1.021 per log-odds increase in the liability to critical infection (roughly equivalent to every 2.72-fold increase in the exposure risk; same below) (MR-IVW, CI: 1.008-1.034; pthres=1e-3). The effect size estimates from other methods were similar but were generally attenuated with larger number of SNPs included. This may be due to weak instruments bias that bias towards the null. This may also be due to ‘winner’s curse’^26^ that overestimates SNP-exposure effects, which in turn biases the MR estimate towards zero. We also observed that liability to hospitalized infection (B2) was associated with ADHD (MR-IVW, OR=1.052, CI: 1.022-1.083; pthres=1e-4).

As for bipolar disorder (BD), we observed primarily that liability to hospitalized infection (B2) was associated with bipolar II disorder (BD-II). The causal effect estimate by MR-IVW was 1.041 (CI:1.013 -1.070, pthres=1e-3). The associations were observed across multiple methods (MR-IVW, MR-Egger, Egger with SIMEX correction, MR-RAPS) and pthres. For SCZ, we observed associations when critical or hospitalized infection were treated as the exposure (analysis A2: MR-IVW, OR=1.049, CI 1.019-1.080, pthres=1e-6; analysis B2, MR-IVW, OR=1.019, CI 1.008-1.030, pthres=1e-2). We also observed evidence that liability to critical COVID-19 was casually associated with PTSD (MR-IVW: OR=1.020, CI 1.009-1.031, pthres=1e-3). We also observed associations of A2 and B2 with suicidal thoughts (ST) by GSMR and SIMEX. Besides, we observed that (genetically predicted) hospitalized infection (B2) and infection in general (C2) may be associated with opioid dependence (Table S1), yet the results did not pass Steiger test of directionality. This indicates the direction of causality may not be reliably inferred, however the significant results still implied genetic overlap between the exposure and outcome.

The results with FDR<0.1 are shown in Table S1. In general similar psychiatric traits were implicated, but more consistent associations were observed (especially with PTSD and ADHD). In addition, bipolar disorder (overall) also showed significant associations.

#### Correlated SNPs as instruments

With correlated SNPs as instruments, as expected, the power to detect associations is generally higher. We shall focus on the results that are significant (with FDR<0.05) across at least two p-value thresholds or two r2-clumping thresholds, for higher robustness. Most of the psychiatric traits/disorders implicated in independent-SNP analysis also showed significant associations with correlated instruments, including BD II, SCZ, PTSD and ST. For BD, we also observed several significant associations with overall BD (Analysis B2: MR-IVW, OR=1.043, CI 1.023-1.063, pthres=1e-4, r2=0.05). Besides, it was observed that liability to critical and hospitalized disease were causally associated with suicidal thoughts across multiple r^2^ thresholds, using MR-Egger (analysis A2, OR=1.034, CI 1.010-1.059, pthres=1e-7, r2=0.15; analysis B2, OR 1.054, CI 1.016-1.094, pthres=1e-7, r2=0.05).

We also observed associations with several other neuropsychiatric disorders not found in independent-SNP analysis. For example, significant associations with OCD and anxiety disorders were detected by MR-Egger; however, we did not observe significant (FDR<0.05) associations using other MR methods. Since we conservatively restrict the number of SNPs to <350 and pthres<=1e-4 to avoid unstable causal estimates, some of the associations in independent-SNP analysis may not be observed in the correlated-SNP analysis.

At a more relaxed FDR threshold of 0.1, we observed several more psychiatric traits being associated with COVID-19 phenotypes (Table S2). Such traits/disorders included BD (overall), BD I, SCZ, ADHD and PTSD, among others. For instance, it was observed that liability to critical and hospitalized COVID-19 infection were causally associated with ADHD (Analysis A2: GSMR, OR=1.024, CI: 1.007-1.042, pthres=1e-4, r2 =0.05; Analysis B2: GSMR, OR=1.030, CI:1.006-1.056; pthres=1e-4, r2=0.05). COVID-19 Infection in general (C2) was also associated with Alzheimer disease (AD) at FDR<0.1 (MR-IVW, OR=1.039, CI: 1.014-1.065, pthres=5e-8, r2=0.05).

#### COVID-19 severity and effects on psychiatric disorders

Overall speaking, most significant causal associations were observed when critical or hospitalized infection was considered as the exposure, suggesting that in general neuropsychiatric sequelae are more likely to be caused by severe rather than mild infections. Only a few phenotypes showed relatively consistent association with C2 (infection in general), such as ADHD and PTSD.

### MR analysis with COVID-19 as outcome and liability to neuropsychiatric disorders as exposure

#### Independent SNPs (r^2^=0.001) as instruments

We observed a smaller number of significant results compared to the case when psychiatric disorders were considered as the outcome. At an FDR threshold of 0.05, liability to ALD, OD and suicide were associated with increased risks or severity of COVID-19 (Table 4). Neuroticism was negatively associated with the disease across all three phenotypes, however they did not pass the Steiger test of directionality, implying that there is genetic overlap but the direction of causality cannot be reliably inferred from the data. At FDR<0.1 (Table S3), similar results were observed but we also found PTSD to be associated with increased risk of C2 (any infection).

#### Correlated SNPs as instruments

We observed more associations with correlated SNPs as instruments, presumably due to better power with larger number of instruments (Table 5). Again we focus on significant (FDR<0.05) exposure-outcome associations across at least two p-value thresholds or two r2-clumping thresholds. Liability to ADHD was causally associated with higher risks of hospitalized or critical infection compared to the general population (analysis A2, MR-IVW; OR=1.112, CI: 1.046-1.182, pthres=1e-4, r2=0.1; analysis B2, MR-Egger; OR=1.136, CI:1.069-1.206, pthres=1e-4, r2=0.05). There was also suggestive evidence (FDR<0.1) that liability to ADHD was associated with higher risks of being test-positive. As for other psychiatric disorders, ALD showed positive association with A2 using MR-Egger (OR=1.209, CI: 1.055-1.386, pthres=1e-4, r2=0.05); association was also observed with C2. Similar to findings from independent SNPs, liability to PTSD was associated with being infected (C2). Several psychiatric traits showed inverse associations with infection phenotypes. Anxious feeling (UKBB data field 1980) and neuroticism were both associated with lower risks of infection or severe illness compared to the population, and the associations were consistent across different methods and p-value thresholds. We also observed that AD was associated with lower risks of critical infection (A2). At FDR<0.1 (Table S4), the results were similar, except that we also observed that MDD was associated with higher risks of infection or severe infection.

### MR analysis with liability to pneumonia as exposure and neuropsychiatric disorders as outcome

Full results for MR analysis with pneumonia can be found in Tables 6 and 7. Based on our MR analysis, we observed different patterns of neuropsychiatric complications of pneumonia, when compared to COVID-19. Figure 1 shows the pattern of significant causal associations in a ‘heatmap’. Interestingly, there is little overlap between COVID-19 and pneumonia in terms of causal associations with neuropsychiatric complications. Overall, liability to pneumonia was casually associated with increased risks/levels of depressive symptoms, MDD, neuroticism, insomnia (at FDR<0.1), anxiety disorders and cannabis use disorder (CUD) (Table S5-S6).

### MR analysis with liability neuropsychiatric disorders as exposure and pneumonia as outcome

Full results for MR analysis with liability to pneumonia as outcome are given in Tables 8 and 9. With independent SNPs as instruments, we observed that the liability to multiple PsyD were casually associated with higher risks of pneumonia across multiple pthres and r2-clumping thresholds. The associated disorders included for example ADHD (OR=1.115, MR-IVW, CI: 1.032-1.205, pthres=5e-8), CUD (OR=1.268, MR-IVW, CI:1.077-1.493, pthres=5e-8), depressive symptoms (beta=0.443, SE=0.143, pthres=1e-6), MDD (OR=1.140, MR-IVW, CI:1.022-1.271, pthres=1e-7) and anxiety disorders (OR=1.047, CI:1.014-1.080, pthres=1e-4). In addition, insomnia and neuroticism were also associated with elevated risk of pneumonia. On the other hand, BD I showed an inverse association with the risk of pneumonia. The above findings were largely consistent using SNPs in LD as instruments (Table 9). Figure 2 showed that the psyD leading to increased risks of pneumonia and COVID-19 were in general different without substantial overlap.

### Genetic correlation (rg) by LD score regression

For genetic correlation analysis, most associations with COVID-19 did not pass multiple testing correction by FDR. Hospitalized COVID-19 infection showed significant rg with CUD (rg=0.340, FDR=2.82e-3), ADHD (rg=0.213, FDR=2.64e-2), and anxiety disorders (rg=0.362, FDR=2.64e-2) at FDR<0.05 (Table 10). Other traits showing at least nominal associations (i.e. p<0.05) with critical/hospitalized illness included psychotic experience (B2, rg=0.543), MDD (A2/B2, rg=0.106 and 0.130 respectively), insomnia (B2, rg=0.126), ADHD (A2, rg=0.146) and BD II (A2, rg = 0.191). Of note, all nominally significant results were related to critical/hospitalized infection (A2 or B2) only. For pneumonia, a large variety of psychiatric traits were found to have significant genetic correlations (Table 10), mostly positive (except OCD). For example, highly significant rg (FDR <1e-4) were observed for MDD, ADHD, insomnia, CUD, anxiety disorders and depressive symptoms.

**Table 10.** Genetic correlation between COVID-19 and Pneumonia with PsyD.

| p1 | p2 | rg | se | z | p | p.adjust |
| --- | --- | --- | --- | --- | --- | --- |
| B2 | adhd2019 | 0.21 | 0.07 | 3.03 | 2.44E-03 | 2.64E-02 |
| B2 | cud.eur.exclude | 0.34 | 0.09 | 3.83 | 1.29E-04 | 2.82E-03 |
| B2 | mdd2019 | 0.13 | 0.05 | 2.50 | 1.26E-02 | 7.98E-02 |
| B2 | psycho.experi | 0.54 | 0.21 | 2.56 | 1.04E-02 | 7.91E-02 |
| B2 | meta.anxiety | 0.36 | 0.12 | 2.99 | 2.78E-03 | 2.64E-02 |
| FP | adhd2019 | 0.46 | 0.07 | 6.12 | 9.28E-10 | 1.64E-08 |
| FP | anx | 0.08 | 0.04 | 2.04 | 4.10E-02 | 6.11E-02 |
| FP | bip.2021.all | 0.14 | 0.05 | 2.91 | 3.60E-03 | 8.32E-03 |
| FP | bip.2021.ii | 0.33 | 0.09 | 3.54 | 4.02E-04 | 1.35E-03 |
| FP | cud_eur | 0.54 | 0.10 | 5.36 | 8.43E-08 | 6.23E-07 |
| FP | cud.eur.exclude | 0.49 | 0.09 | 5.80 | 6.79E-09 | 6.28E-08 |
| FP | dep.sym2016 | 0.38 | 0.08 | 4.50 | 6.84E-06 | 3.62E-05 |
| FP | ever.sh | 0.41 | 0.14 | 2.94 | 3.28E-03 | 8.08E-03 |
| FP | insom2019 | 0.39 | 0.06 | 6.06 | 1.33E-09 | 1.64E-08 |
| FP | mdd.2021.broad | 0.21 | 0.10 | 2.09 | 3.64E-02 | 6.11E-02 |
| FP | mdd.2021.narrow | 0.26 | 0.12 | 2.23 | 2.56E-02 | 4.73E-02 |
| FP | mdd2019 | 0.32 | 0.05 | 6.34 | 2.29E-10 | 8.47E-09 |
| FP | neuroti_eur_ukbb_2018 | 0.15 | 0.05 | 2.88 | 3.96E-03 | 8.61E-03 |
| FP | neuroti.meta2016 | 0.18 | 0.07 | 2.58 | 9.83E-03 | 1.91E-02 |
| FP | ocd2018 | -0.23 | 0.11 | -2.01 | 4.49E-02 | 6.39E-02 |
| FP | ptsd2019 | 0.51 | 0.12 | 4.11 | 3.97E-05 | 1.63E-04 |
| FP | sa.ipsych | 0.43 | 0.21 | 2.06 | 3.96E-02 | 6.11E-02 |
| FP | scz.wave3.v2 | 0.10 | 0.05 | 2.04 | 4.13E-02 | 6.11E-02 |
| FP | st | 0.79 | 0.19 | 4.17 | 3.04E-05 | 1.41E-04 |
| FP | st.dep | 0.55 | 0.18 | 3.16 | 1.59E-03 | 4.20E-03 |
| FP | alco2018.beta | 0.57 | 0.14 | 3.98 | 6.76E-05 | 2.50E-04 |
| FP | meta.anxiety | 0.54 | 0.11 | 5.18 | 2.24E-07 | 1.38E-06 |
| FP | od.case_unexposed.eur | 0.46 | 0.21 | 2.20 | 2.76E-02 | 4.87E-02 |
| FP | od.case_unexposed.trans | 0.55 | 0.20 | 2.76 | 5.71E-03 | 1.17E-02 |

\*Represents results with 0.05< FDR-adjusted $p$ <0.1. All other results have FDR-adjusted $p$ <0.05.

## Discussion

### Overview

Overall speaking, we observed potential bi-directional causal associations between neuropsychiatric disorders and COVID-19 (including severe illness). We observed that liability to COVID-19 (especially critical or severe illness requiring hospitalization) may be causally linked to ADHD, BD (especially BD II), PTSD, SCZ and ST. Conversely, liability to a few psychiatric traits/disorders, for example ADHD, ALD, OD, PTSD and SA may be causally associated with higher risks of COVID-19 infection or severe disease. Interestingly, the patterns of causal associations with psychiatric traits appeared to be different when we compared COVID-19 to pneumonia.

### Neuropsychiatric disorders as sequelae to COVID-19 infection

The current MR study provides support that COVID-19 may be casually linked to a number of neuropsychiatric sequelae. A few studies have attempted to examine neuropsychiatric consequences of COVID-19 to date, which will be highlighted below.

One of the largest observational studies was performed by Taquet et al.^4^, who observed increased risks of mood disorders, anxiety disorders, insomnia and dementia as first psychiatric diagnoses within 3 months after the infection in a retrospective cohort. Psychotic disorders were also observed to be of higher incidence after infection if both new and recurrent diagnoses were counted. Based on further details provided in supplementary information (Table S8)^4^, compared to influenza, the risks of bipolar disorder, depressive episode and PTSD all appeared to be increased in COVID-19 patients, with the latter two being statistically significant. A very recent further analysis^29^ with a larger sample (*N*=236379) and longer (6-month) follow-up showed similar findings. Dementia, mood, anxiety, psychotic and substance use disorders were all significantly associated with COVID-19 infection compared to influenza or other respiratory tract infections, and the effects were generally larger for more severe cases.

These findings were largely consistent with our current MR analyses. Our results using MR suggests that the associations with some of the above psychiatric traits/disorders may be causal, and may not be fully explained by confounding factors alone. As stated by the authors, the previous study^4^ was limited by possibility of (residual) confounding factors; in addition, socioeconomic factors, which is known to be associated with both infection risk/severity^30–34^ and some other psychiatric disorders, were not modelled. It is also possible that some disorders (e.g. dementia) was undiagnosed but were detected during follow-up after the infection as a result of detailed assessments and monitoring for mental health problems, hence the disorder may not be a consequence of infection. Another related possible bias is that COVID-19 patients may have received more attention and follow-ups for mental health issues compared to flu or other RTI patients. The MR approach substantially reduces the risk of confounding (e.g. by socioeconomic status) or uncertainty about temporal sequence of events, and avoids possible bias of differential assessment after COVID-19 vs other comparison diseases.

With respect to specific disorders implicated in our MR study, we observed relatively consistent evidence of causal links with PTSD (especially at FDR<0.1). Several studies have reported on increased incidence of PTSD after COVID-19 infection. For example, it was reported that up to ∼30% of patients developed PTSD after an acute infection^35, 36^. Other studies have also looked into the risk factors and clinical correlates of PTSD^37^, showing that more severe disease may be associated with higher risks of PTSD.

BD (especially BD II) and SCZ were also implicated in our analysis as possible sequelae of infection. Tanquet et al.^4^ reported that the incidence of both psychotic and mood disorders were significantly higher than influenza and other RTI from a US population, although data specifically on BD II and schizophrenia were not available. There were also a number of case reports on psychosis following COVID-19 infection^38, 39^, but longer follow-up is required to delineate the course of the illness. However, there are otherwise very few studies on whether new-onset or recurrent BD and SCZ is a consequence of COVID-19, given that both disorders are of relatively low incidence and the available follow-up period is short.

As for suicidal thoughts (ST), as COVID-19 may be associated with a range of psychiatric disorders and physical symptoms, it has been suggested that there may be increased suicidal risk among COVID-19 survivors^40, 41^. A recent study on veterans showed^42^ increased suicidal ideations in infected subjects. The possible mechanisms underlying heightened suicidal risks was summarized in a recent review^43^.

For ADHD, although ADHD is generally a childhood-onset disorder, the current analysis may also be considered to reflect the effect of the exposure on the propensity to ADHD or the continuum of ADHD symptoms^44^. Of note, ADHD *symptoms* such as inattention and poor concentration are known to be associated with coronavirus infections^36^ and are considered part of the ‘long-COVID’ syndrome^27^. The current MR analysis provides some support that COVID-19 may be causally related to ADHD symptoms. Finally, for OD, a recent study has shown that opioid analgesics are significantly more commonly received by COVID-19 survivors (HR=9.39) ^28^, which may be related to chronic pain symptoms.

#### Neuropsychiatric disorders as sequelae to pneumonia

We found that pneumonia may be a causal risk factor for depressive symptoms, MDD, neuroticism, insomnia (at FDR<0.1), anxiety disorders and CUD. It was reported that hospitalization for pneumonia was associated with higher odds of substantial depressive symptoms (OR = 1.63)^45^. However, there were few studies on the neuropsychiatric sequelae of pneumonia, which precludes a detailed comparison of the current findings to previous studies.

A number of studies have investigated whether psychiatric disorders may increase the risk or severity of COVID-19 infection. For example, Yang et al^46^ showed that in general pre-existing psychiatric disorders were associated with higher risks of COVID-19 infection and mortality, based on the UKBB sample. Considering individual disorders, depression, anxiety, substance misuse and psychotic disorders were all associated at least one of the outcomes (COVID-19 infection, hospitalized infection or fatality)^46^ in the above study. However, the number of cases for stress-related disorders and some other disorders (e.g. psychosis) is relatively small, and the UKBB may not be representative of the underlying population due to ‘healthy volunteer bias’^47^. Tanquet et al. also reported that patients with a psychiatric diagnosis within a year of the COVID-19 outbreak had ∼65% higher risk of being infected compared to a matched cohort with matched physical risk factors. However, associations of individual psychiatric disorders with infection were not reported in the study. In another US study of electronic health records^1^, ADHD, BD, MDD and SCZ were all associated with elevated risks of infection. On the other hand, Lee et al.^2^ did not find an association of mental illness with infection but there was a modest association (adjusted OR of 1.27) with severity of infection. One important limitation of observational studies is the risk of residual confounding. For example, socioeconomic status is not controlled for by Tanquet et al., and incomplete data on some covariates may lead to inadequate control for confounders. In addition, the above studies typically focused on a few psychiatric disorders, and the coverage is not as comprehensive as the current study.

In this MR study, liability to AD, ADHD, ALD, OD, PTSD and SA were associated with increased risk and/or severity of COVID-19 infection/severity. As discussed above, there was evidence that ADHD^1^ and substance use disorders^46^ may be associated with heightened risk of COVID-19. There were no previous studies that directly addressed association of PTSD and history of suicide with susceptibility to infection. However, in Yang et al^46^, stress-related disorders were associated with higher risks of infection, hospitalization and mortality, although the results were non-significant. Suicide attempts (SA) is linked to many psychiatric disorders (e.g. mood, anxiety and psychotic disorders) and previous studies have shown that prior psychiatric disorders as a whole were associated with COVID-19.

There have been relatively few studies on the associations of psychiatric disorders with risks/severity of pneumonia. A study using linked hospital records showed that the risk of pneumococcal disease was elevated for patients with SCZ, BD, anxiety or depression^8^. We found potential causal relationships of anxiety and depression phenotypes with pneumonia in this study. SCZ showed positive associations with pneumonia in MR-Egger analysis with correlated SNPs, but the association was less consistent than that observed for anxiety or depression. Unexpectedly, we observed negative associations of BD I with pneumonia, contrary to the finding of the above study. The underlying reason is unknown, but could be due to differences in the phenotype under study (BD in general vs BD I), heterogeneity of the study samples, differences in comorbidity patterns etc. A previous study has shown that lithium (which is commonly prescribed in BD) has protective effects against pneumonia^49^, however antipsychotics may have the opposite effects (cite) so it is hard to conclude the direction of effect.

Another study showed that depression is associated with heightened risks of ICU admission, mechanical ventilation and mortality from pneumonia^50^. As for ADHD, a German study using claims data^51^ revealed that ADHD may be associated with higher risks of multiple comorbidities including viral pneumonia. Another interesting finding was that cannabis use disorders was consistently associated with higher risk of pneumonia by MR. It has been proposed that cannabis is linked to various lung pathologies and may weaken the immune response thus raising the risk of pneumonia^52, 53^. Another case-control study^54^ showed that regular cannabis use was associated with higher pneumonia risks, regardless of tobacco co-use.

A recent bi-directional MR study by Luykx and Lin found that liability to BD and SCZ combined together (based on GWAS meta-analysis of BD and SCZ) may be associated with increased risk of COVID-19 infection^30^. However, the study did not reveal any other significant causal associations, either considering COVID-19 as outcome or exposure. Our reported findings are partially different which can be due to various reasons. For example, Luykx and Lin considered a stringent p-value threshold of 5e-8 and hence only a few instruments were included in each analysis, especially when COVID-19 phenotypes were considered as the exposure, which may lead to limited statistical power. Here we have employed more relaxed p-value thresholds, which have shown by several previous studies and our own simulations to maintain type I error control. Inclusion of larger number of instrumental SNPs in LD may also increase the power. In addition, we have also covered a much wider range of psychiatric traits/disorders than the previous study, and a number of psychiatric GWAS datasets we used were also different or more updated (hence of larger sample sizes) (see Table 1). We also included pneumonia as an outcome and exposure; to our knowledge, no previous studies have investigated causal relationship of psychiatric disorders with pneumonia.

## Strengths and limitations

There are several limitations of this study. We have employed the latest and GWAS summary statistics to date for COVID-19, however heterogeneity (e.g. in disease severity, demographics) may be present across different constituent studies. It is also difficult to verify that the reason for hospitalization is due to (severe) COVID-19 symptoms, and the criteria for admission may differ across cohorts. The control population is composed of the general population, and asymptomatic patients and patients with mild symptoms may be missed. The sample sizes of severe and critical cases were still relatively modest, hence the statistical power to detect weaker associations may be limited. Similar limitations, such as heterogeneity across studies and modest sample sizes, are also present for many of the neuropsychiatric GWAS datasets. For example, the severity and presentation of MDD can be highly heterogeneous, yet they may be grouped under the same category. We have tried to include more well-characterized phenotypes (e.g. melancholic depression, severe MDD requiring ECT etc.), but such studies are usually of smaller sample sizes.

Many of our findings are supported by previous studies. However, for some associations supported by the literature, we were unable to verify their causal relationship in this MR study. For example, increased risks of depression and anxiety disorders after COVID-19 have been reported in observational studies^4^, but our MR analysis did not support this observation. A number of possible reasons may explain this. As discussed above, statistical power may be insufficient to detect modest (causal) associations due to limited sample sizes of COVID-19 and/or the psychiatric GWAS datasets. Heterogeneity in inclusion/exclusion criteria and sample characteristics or definition of psychiatric outcomes may also explain the differences. On the other hand, it is possible that previously observed associations may be (partially) explained by confounding factors. For example, low socioeconomic status (SES) and poor physical health are linked to both COVID-19 infection and depression/anxiety disorders. Some confounders may remain uncontrolled for or not adequately controlled for.

There are other limitations of the MR methodology. For more detailed discussions of the strengths and shortcomings of MR, please also refer to other papers^20, 55–57^. For example, horizontal pleiotropy (the instrumental SNP being associated with the outcome via another pathway not through the exposure) may affect the validity of results, which can be accounted for by some MR methods^58, 59^. Yet different MR method require different assumptions such as the InSIDE assumption^60^ for MR-Egger, systematic and idiosyncratic pleiotropy for MR-RAPS and so on. We therefore have attempted multiple methods and results robust across different approaches may be more reliable for further studies. We also note that there is sample overlap between the pneumonia GWAS datasets and some other neuropsychiatric datasets (both included UKBB samples); as such results may bias towards the confounded estimate and should be interpreted with caution. However, we expect the bias to be relatively small as instrument strengths were generally strong. Also, the primary focus of this study is on COVID-19, and we have used COVID-19 summary statistics that did not contain UKBB samples.

## Conclusions

To our knowledge, this is the most comprehensive MR study to date investigating *bi-directional* causal relationships between COVID-19 and neuropsychiatric disorders/traits. This is also the first MR study of neuropsychiatric disorders with pneumonia. We found tentative evidence of causal links between several psychiatric traits/disorders and COVID-19, especially for severe infections. The patterns of associations appeared to be different when compared to pneumonia. Further replications in larger and prospective cohorts are required, and the underlying mechanisms require further studies.

## Data Availability

All GWAS summary data are publicly available online as described in the paper.

## Acknowledgements

This work was supported partially by a National Natural Science Foundation China (NSFC) grant (81971706), Lo Kwee Seong Biomedical Research Fund from The Chinese University of Hong Kong (CUHK) and a Direct Grant from CUHK. We thank Mr. Kenneth C.Y. Wong for preparation of some GWAS summary datasets.

## Author Contribution

Conception and design: HCS (lead), with input from YX and SR. Study supervision: HCS. Funding acquisition: HCS. Methodology: HCS (lead), YX. Data curation: YX, JQ, RZ, CKLC, SR. Data analysis: YX (lead), JQ, RZ, SR. Data interpretation: HCS, YX. Preparation of first draft of manuscript: HCS (lead), YX.

## Supplementary Information

All supplementary Tables and notes are available at the journal’s website and at https://drive.google.com/drive/folders/18ZECXMeoDL1NRpweKsD0wYWGKm_y4QPa?usp=sharing

